# Kinetics of SARS-CoV-2 infection in the human upper and lower respiratory tracts and their relationship with infectiousness

**DOI:** 10.1101/2020.09.25.20201772

**Authors:** Ruian Ke, Carolin Zitzmann, Ruy M. Ribeiro, Alan S. Perelson

## Abstract

SARS-CoV-2 is a human pathogen that causes infection in both the upper respiratory tract (URT) and the lower respiratory tract (LRT). The viral kinetics of SARS-CoV-2 infection and how they relate to infectiousness and disease progression are not well understood. Here, we develop data-driven viral dynamic models of SARS-CoV-2 infection in both the URT and LRT. We fit the models to viral load data from patients with likely infection dates known, we estimated that infected individuals with a longer incubation period had lower rates of viral growth, took longer to reach peak viremia in the URT, and had higher chances of presymptomatic transmission. We then developed a model linking viral load to infectiousness. We found that to explain the substantial fraction of transmissions occurring presymptomatically, a person’s infectiousness should depend on a saturating function of the viral load, making the logarithm of the URT viral load a better surrogate of infectiousness than the viral load itself. Comparing the roles of target-cell limitation, the innate immune response, proliferation of target cells and spatial infection in the LRT, we found that spatial dissemination in the lungs is likely to be an important process in sustaining the prolonged high viral loads. Overall, our models provide a quantitative framework for predicting how SARS-CoV-2 within-host dynamics determine infectiousness and represent a step towards quantifying how viral load dynamics and the immune responses determine disease severity.

**Significance:** A quantitative understanding of the kinetics of SARS-CoV-2 infection is key to understanding the development of infectiousness and disease symptoms. To address this need, we developed data-driven within-host models of SARS-CoV-2 infection and showed that lower rates of viral growth lead to longer incubation periods and higher chances of presymptomatic transmission. We found that the logarithm of the URT viral load serves an appropriate surrogate for a person’s infectiousness. We then developed a mechanistic model for infectiousness and showed that a saturation effect in the dependence of transmission on viral load gives rise to this relationship. We also provide evidence of spatial dissemination in the lungs as an important process in sustaining prolonged high viral loads in the LRT.

## Introduction

SARS-CoV-2 is a new human pathogen that causes COVID-19 (1). It is highly contagious and spread rapidly across the globe. It is estimated that the outbreak grew extremely fast and doubled every 2.3-3.6 days in the absence of strong control measures (2-4) and it has caused more than 900,000 deaths worldwide as of September 2020. Extensive efforts to develop effective treatments and vaccines are underway.

SARS-CoV-2 infects cells in both the upper respiratory tract (URT) and the lower respiratory tract (LRT) (5). It enters host cells via the receptor ACE-2 (angiotensin converting enzyme 2) on epithelial cell surfaces (6). Structural analysis suggests that SARS-CoV-2 binds to the receptor >10-fold more efficiently than SARS-CoV-1 (7), partially explaining the comparatively high contagiousness of the virus. It is likely that the ability of the virus to effectively infect cells in the URT allows the virus to be transmitted before symptom onset (8, 9), an important factor that makes the virus difficult to control (10). It has been suggested infectiousness is positively related to viral load in the URT (11); however, it is not clear how these are quantitatively related. For example, both viral load and the logarithm of viral load have been used as surrogates of infectiousness (12). A quantitative understanding of the relationship is lacking. Quantifying it is critical because it would allow for more precise predictions of the infectiousness of certain groups of infected individuals, such as children and asymptomatic individuals, based on their viral load measurements (13, 14). This would in turn help to better inform policy decisions.

The pathogenesis of COVID-19 is not fully understood. Recent data shows that SARS-CoV-2 viral load in the LRT correlated with lung damage and disease severity (15, 16). SARS-CoV-2 infection in the LRT can lead to excessive production of proinflammatory molecules and an exacerbated inflammatory response (17), which can cause lesions, damage in the lungs and severe pneumonia (16, 18-20). A steroid, dexamethasone, that dampens inflammatory responses, has shown clinical benefit in ventilated patients in the RECOVERY trial (21). Some reports suggested that high viral loads are associated with a higher risk of disease severity and a higher rate of mortality in adults (22-24), while others found that viral load is higher in children than in adults (25). Quantifying the viral dynamics in the LRT will help to better understand these issues.

Mathematical modeling plays a crucial role in understanding the pathogenesis of viral diseases and in the development of effective therapeutics and treatment strategies for viruses such as HIV, hepatitis C, influenza, Zika and Ebola (26-31). Mathematical modeling has been applied, by us and others (32-36), to understand SARS-CoV-2 infection in hospitalized patients and the potential impact of therapy. However, these studies modeled a single physiological compartment even though SARS-CoV-2 infects both the URT and the LRT and there were large uncertainties in the parameter estimates because the patient infection dates were unknown. Here, we develop within-host models of SARS-CoV-2 infection in both the URT and the LRT. We fit the models to a set of viral load data collected from the first cluster of individuals in Germany, for whom infection dates were reported (5, 37). We analyze how the onset of symptoms is affected by viral dynamics, and the potential mechanisms for the prolonged period of detectable virus in the LRT observed in clinical studies (5, 16). We also analyzed the relationship between viral load and potential infectiousness of a person. Using existing epidemiological evidence, we developed and constrained a probabilistic model to quantify this relationship. Lastly, we develop several models to explain the prolonged period of virus infection in the LRT and explore how treatments may impact virus transmissibility and disease outcomes.

## Results

### Dynamics of early infection

As in previous work (33, 35), we first constructed a target cell limited (TCL) model (see Methods) to quantify the early dynamics of SARS-CoV-2 infection in both the URT and the LRT. We used this model to estimate key parameters, such as the time to peak viremia and the within-host basic reproductive number (R_0_) and their relationships with the time from infection to symptom onset, i.e. the incubation period, which is known for each patient in the dataset we analyzed (37). We fit the model to data collected during the first 14 days of infection. Beyond this time, the effect of an adaptive immune response may be important (38), and we model that separately (see below). Further, regeneration of target cells by proliferation may become significant especially in the LRT (19, 39) and we consider a model later that incorporates proliferation in the LRT (see below) as virus in the URT has declined to undetectable or almost undetectable levels by this time (Fig. 1). During this early period, the dynamics are typical of a target cell limited acute respiratory infection, i.e. viral loads measured in both throat swabs and sputum samples increased to a viral peak and declined afterwards, and are relatively homogeneous across patients (Fig. 1). In one patient (Patient 16 as numbered in Ref. (37)), the first data point is at day 15 post infection. Therefore, this patient was excluded from these initial fits. We used a nonlinear mixed effect modeling approach to fit data collected (URT and LRT) from the other 8 patients simultaneously (see Methods and Supplementary Material).

**Figure 1.**
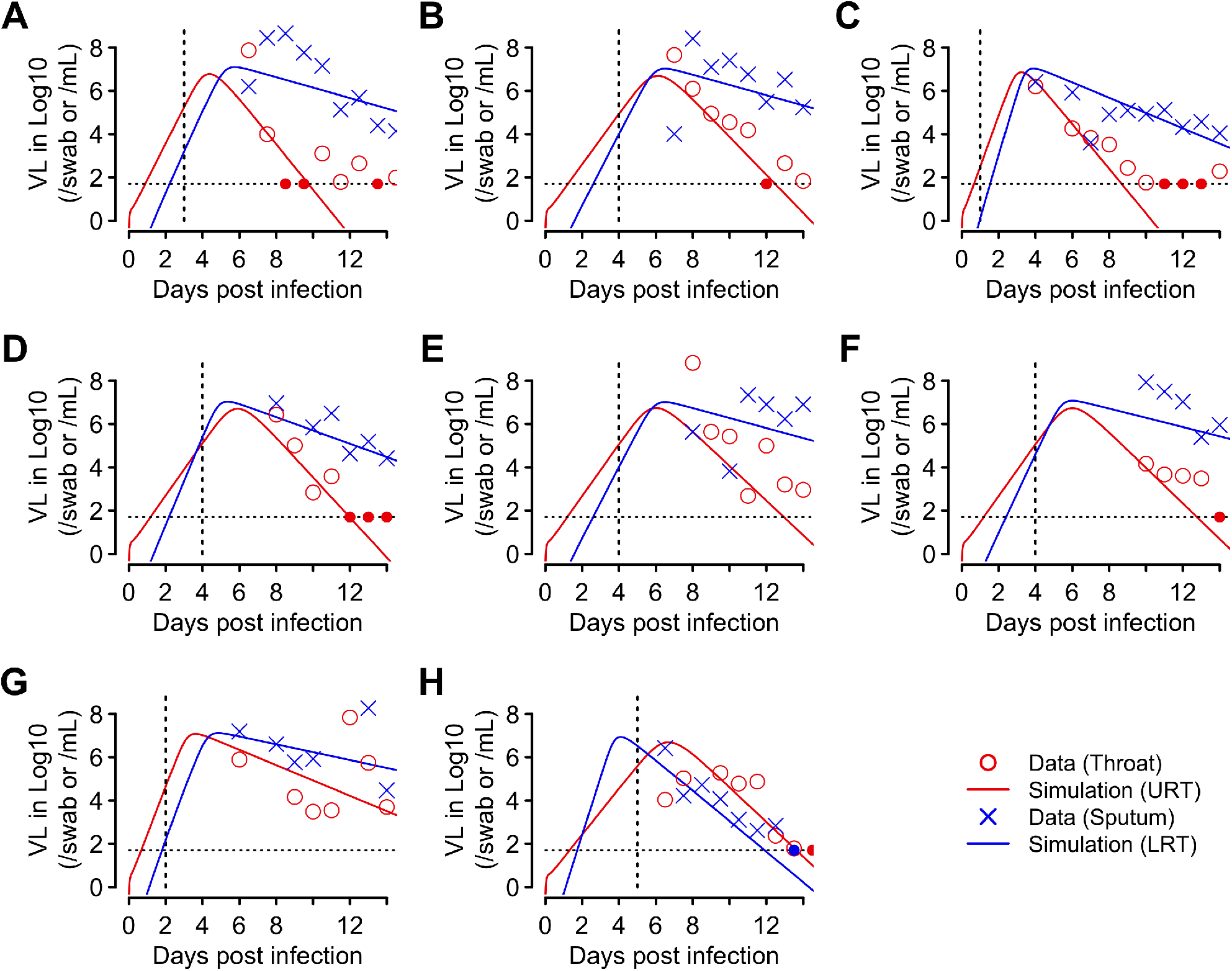
The fit of the target cell limited model (Eqns. 2 and 3) to the early viral kinetics from the URT and the LRT. The model (lines) was simulated using the best-fit individual parameter values estimated by a non-linear mixed effect modeling approach (Table S2). Red and blue denote viral load kinetics in the URT and the LRT, respectively. Symbols show the data from throat swabs (red circles) and the sputum samples (blue ‘x’s) as reported in Ref. (5). Vertical dashed black lines denote the time of symptom onset for each individual as reported in Ref. (37). Horizontal dashed black lines show the limit of detection, and filled circles indicate data points below the limit of detection.

To understand whether the duration of the incubation period is related to parameters governing the viral dynamics, we fitted the model allowing for the duration of the incubation period to be a covariate for each of the fitted parameters (see Methods and Supplementary Material). This leads to 7 model variants (Table S1). Comparing these models using the Akaike Information Criteria (AIC), we found that the best ones are those that assume the infectivity of the virus, *β*_*T*_, or the virus production rate, *π*_*T*_, in the URT vary inversely with the duration of the incubation period (Table S1)

The best model, in which *β*_*T*_ varies inversely with the incubation period, fits the data from both the URT and the LRT (Fig. 1, and see Table 1 and S2 for the best-fit population parameter values and the individual parameter values, respectively). We estimated that the viral load peaks on average 5.2 days (standard deviation: ±1.3 days) and 5.4 days (±1 day) post infection in the URT and the LRT, respectively, and on average 2 days (±0.2 day) and 2.1 days (±1.2 days) post symptom onset (Table 2). The within-host basic reproductive numbers were estimated to be on average 8.5 (±7.1) and 27.5 (±8.4) in the URT and LRT, respectively (Table 2). Examining the viral dynamic characteristics in the URT and the LRT, we found a significant correlation between *R*_0,*URT*_ and *R*_0,*URT*_ (p-value: 0.003; Fig. S1A), i.e., a high within-host reproductive number in the URT predicts a high reproductive number in the LRT. On the other hand, we found no significant correlation between the URT and the LRT in the time from infection to the viral load peak, nor in the time from symptom onset to the viral load peak (Figs. S1B and S1C).

**Table 1.** Estimated population parameter values obtained by fitting the TCL model to the early viral kinetic data collected up to 14 days post infection.

| Parameter | Description | Estimated value<br>(population estimate) |
| --- | --- | --- |
| $\beta_T$ | Infectivity parameter (URT) | $1.9 \times 10^{-6}$ swab/day |
| $\sigma_\beta$ | Covariate coefficient of time to symptom onset on $\beta_T$ | -0.33 |
| $\delta$ | Death rate of infected cells (URT) | 1.9 /day |
| $\pi_T$ | Composite parameter for virus production and sampling (URT) | 51.4 /swab/day |
| $\beta_S$ | Infectivity parameter (LRT) | $6.6 \times 10^{-7}$ mL/day |
| $\delta_2$ | Death rate of infected cells (LRT) | 0.63 /day |
| $\pi_S$ | Composite parameter for virus production and sampling (LRT) | 0.35 /mL/day |

**Table 2.** Early viral dynamic characteristics in each infected individual. The characteristics are summarized from simulations using the best-fit individual parameter values of the target cell limited model to the early viral kinetic data (i.e. data collected up to 14 days post infection). SD – standard deviation.

| Panel (ID)* | Incubation period* | Time from infection to VL peak (days) | | Time from symptom onset to VL peak (days) | | $R_{0,URT}$ | $R_{0,LRT}$ |
| --- | --- | --- | --- | --- | --- | --- | --- |
|  |  | URT | LRT | URT | LRT |  |  |
| **A (1)** | 2.5 | 4.4 | 5.7 | 1.9 | 3.2 | 7 | 24.6 |
| **B (2)** | 4 | 6.1 | 6.5 | 2.1 | 2.5 | 4.6 | 21.2 |
| **C (3)** | 1 | 3.2 | 3.9 | 2.2 | 2.9 | 11.9 | 38.9 |
| **D (4)** | 4 | 5.9 | 5.4 | 1.9 | 1.4 | 4.6 | 24.3 |
| **E (7)** | 4 | 6 | 6.5 | 2 | 2.5 | 5.2 | 21.5 |
| **F (8)** | 4 | 6 | 6 | 2 | 2 | 5.1 | 27.4 |
| **G (10)** | 2 | 3.6 | 4.9 | 1.6 | 2.9 | 25 | 42.1 |
| **H (14)** | 4.5 | 6.7 | 4.1 | 2.2 | -0.4 | 4.3 | 19.8 |
| **Mean** | **3.2** | **5.2** | **5.4** | **2** | **2.1** | **8.5** | **27.5** |
| **SD** | **1.3** | **1.3** | **1.0** | **0.2** | **1.2** | **7.1** | **8.4** |

\* Panels correspond to the panels shown in Fig. 1. The ID and the incubation period for each individual are taken from Ref. (37). Viral load is abbreviated as VL.

Next, we performed sensitivity analyses to test how robust our estimates are with respect to variations in the fixed parameter values of the model (Table 3). We varied each of the fixed parameter values in the ranges shown in Table 3 and then re-fit the model to the data. Across the scenarios examined, the estimates of the death rate of infected cells in the URT and the LRT are robust at approximately 2 d^-1^ and 0.7 d^-1^, respectively (Table S3). The within-host reproductive number in the URT *R*_0,*URT*_ ranges between 5-15 (Table S4). The estimated within-host reproductive number in the LRT, *R*_0,*URT*_, spans a wider range and it is mostly affected by variations in the rate of virus transport from the URT to the LRT, Γ (Table S4). This parameter determines how quickly viruses are seeded in the LRT. The lack of knowledge of this parameter means that *R*_0,*URT*_ cannot be reliability determined. If seeding occurs quickly (Γ=0.1/day) then *R*_*0,LRT*_ is similar to *R*_*0,URT*_; on the other hand if this seeding takes longer to occur (Γ=0.001/day) then *R*_*0,LRT*_ will be much larger, since there is less time for the viral load to reach the high levels observed. Overall, the estimated parameters and viral dynamic characteristics are robust against variations in the fixed parameters, with exception for the infectivity and the reproductive number in the LRT, *R*_0,*URT*_ (Tables S3 and S4).

**Table 3.** The fixed parameters in the model and their values.

| **Parameter** | **Description** | **Values** | **Values tested in sensitivity analyses** |
| --- | --- | --- | --- |
| $T_{1,0}$ | Total number of (infection free) target cells in the URT | $4 \times 10^6$ cells | NA |
| $T_{0,2}$ | Total number of (infection free) target cells in the LRT | $4.8 \times 10^8$ cells | NA |
| $I_0$ | Initial number of infected cells in the URT | 1 cell | 10 cells |
| $c$ | Virus clearance rate | 10/day | 5 and 20/day |
| $k$ | 1/k is the eclipse period | 4/day | 3 and 6/day |
| $\Gamma$ | Virus transport rate from the URT to the LRT in the simplified model | 0.01 swab/mL/day | 0.001, 0.1/day |

### Determinants of the duration of the incubation period and infectiousness

Since this model provides a good description of the viral load dynamics, we next asked how these relate to several important epidemiological parameters. Using the best-fit individual parameter values in Table S2, we examined the relationship between the incubation period and characteristics of the viral dynamics. We found that the time from infection to peak viral load in the URT strongly correlates with the incubation period (r^2^=0.98); whereas this duration in the LRT does not (Fig. S2A and B). The within-host reproductive numbers in both the URT and the LRT negatively correlate with the incubation period (Fig. S2C and D). These results suggest that the development of initial symptoms may be associated with how quickly the viral load reaches a high level in the URT and the rates of viral growth in the URT and the LRT.

We next examined how infectiousness is related to the viral load dynamics, since transmission is likely to depend on viral load in the URT (11, 40). Epidemiological studies have shown that infectiousness starts several days before symptom onset and a sizable fraction of transmissions (>30%) occurs presymptomatically (8, 9). We first examined two commonly used measures to summarize the viral load dynamics in the URT (12), the area under the viral load curve (AUC) and the area under the log_10_ of the viral load curve (AUClog). Irrespective of the measure used, we found that the longer the incubation period, the larger the fraction of presymptomatic viral load (Fig. 2; p-values<0.001 for both measures), and thus the more likely presymptomatic transmission occurs.

**Figure 2.**
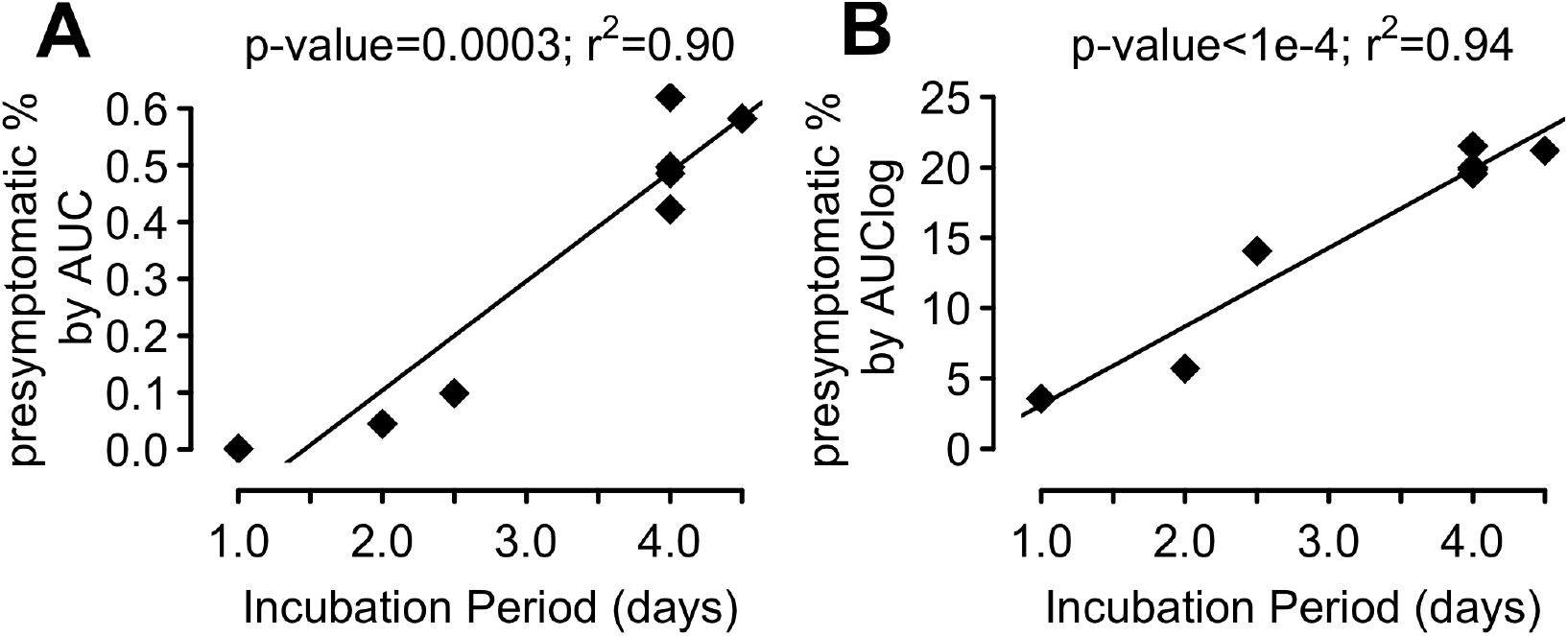
Regression analyses of the relationship between the duration of the incubation period and estimated presymptomatic area under the curve (AUC) percentages for (A) the viral load and (B) the log_**10**_ viral load in the URT.

The fraction of the viral load AUC that occurs during the presymptomatic period, i.e. the “presymptomatic %AUC”, ranges between 0.001% and 0.5% across the 8 patients (Fig. 2A). This suggests that the viral load AUC is unlikely to be a good measure of infectiousness; otherwise, these results would predict that a very small fraction of transmissions occur before symptom onset. In contrast, the presymptomatic %AUClog ranges between 3% and 21% (Fig. 2B), i.e. near the lower bound estimate in Ref. (9). Therefore, the logarithm of viral load, and its corresponding AUC, i.e. AUClog, serves as a better surrogate for infectiousness than the viral load and its corresponding AUC. Note that it is expected that the presymptomatic fraction we calculated for this group of patients is lower than the fraction in the general population, due to the relatively short incubation periods of the patients in our analysis (ranging between 1 and 4.5 days). In the general population, patients with an incubation period longer than 4.5 days are likely to have higher probabilities of presymptomatic transmission (Fig. 2B), and thus would lead to a higher fraction of presymptomatic transmission than the model predicted for the 8 patients in this study.

### Probability of transmission

Next, we sought to provide a physiological basis for the relationship between the logarithm of viral load and infectiousness. We developed a probabilistic model that incorporates the various steps from viral shedding to establishment of infection (see Methods for details). In this model, we define the infectiousness as the probability that an infected person will generate one or more viral particles leading to a successful transmission event for a typical contact of a relatively short duration (minutes to a couple of hours). This probability is expressed as:

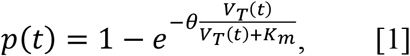

where *V*_*T*_(*t*) is the viral load in a swab taken at time *t* post-infection. We used a Michaelis-Menten term to describe the dependence of the number of viruses shed by a host on their URT viral load. The mathematical form of Eqn. [1] is similar to previous models describing infectiousness of individuals infected by human immunodeficiency virus or influenza as a function of their viral loads (12, 41, 42). Moreover, the Michaelis-Menten term in our model is motivated by experimental results from seasonal coronavirus showing the amount of exhaled virus stops increasing when viral load becomes very large (11) (see Fig. S3 and Methods). That is, exhaled virus is a saturating function of viral load, and *K*_*m*_ is a saturation constant that defines the viral load of half-maximal shedding. This functional form also encompasses a linear dependence on viral load when *K*_*m*_ is very large. *θ* is a constant such that the maximum transmission probability, i.e. the maximum infectiousness, is 1 – *e*^−*θ*^. We set *θ* = 0.05, leading to a maximum transmission probability of approximately 5% for a typical contact of a relatively short duration (Fig. 3). This is generally consistent with the low secondary attack rates per contact reported in multiple studies analyzing contact tracing data (43-45). Note that because *θ* is small and 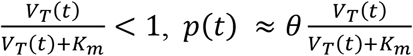. Thus, *K*_*m*_ also corresponds approximately to the viral load at which the infectiousness of an infected person is 50% of the its maximum infectiousness (Fig. 3I).

**Figure 3.**
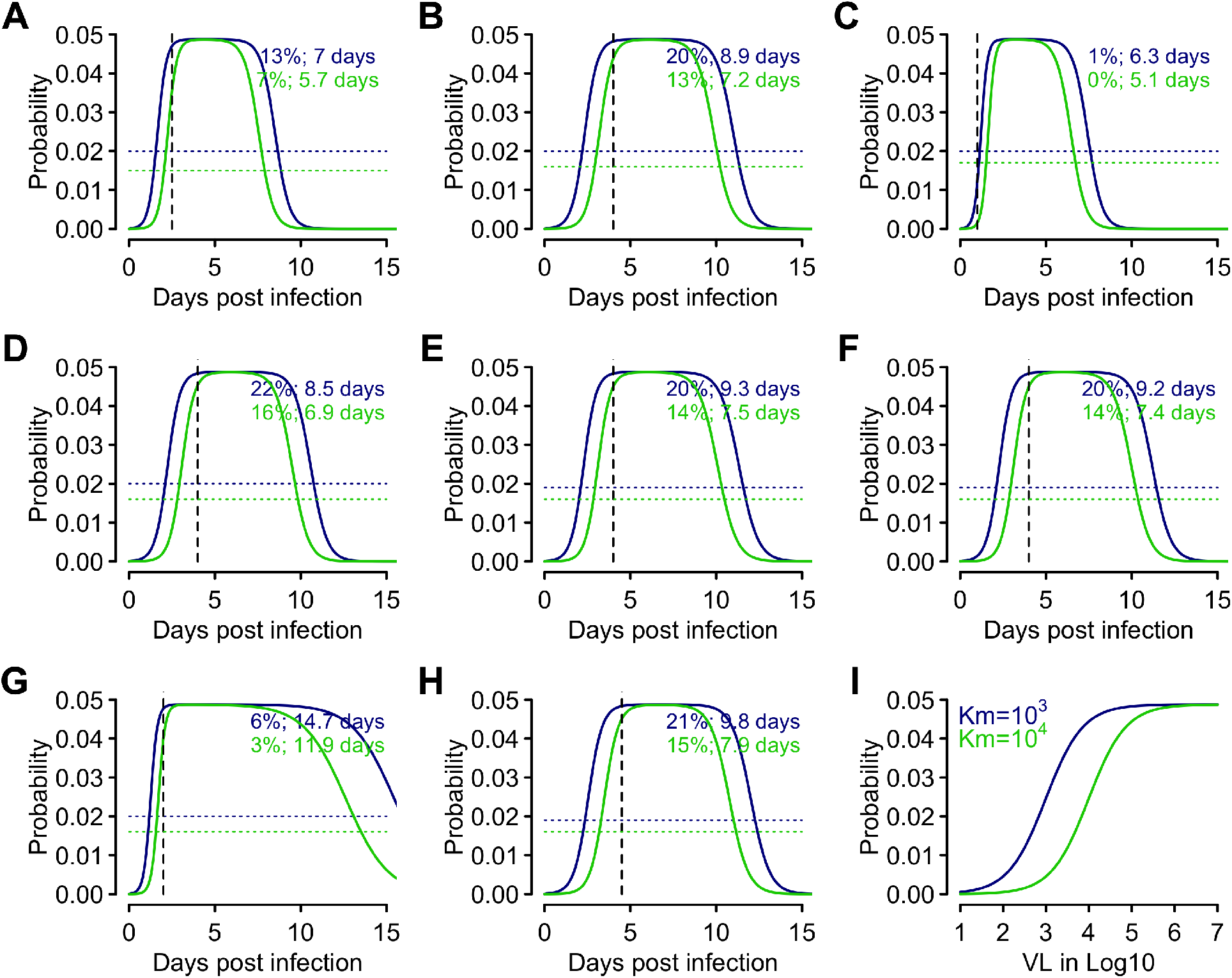
The infectiousness profile predicted by the probability model. **(A-H)** The probability of transmission for a typical contact, i.e. infectiousness, over time predicted for each individual using the model linking viral load to infectiousness. We set *θ* = 0.05 and *K*_*m*_ = 10^3^ (dark blue) or 10^4^ (green) RNA copies. Vertical black lines show the day of symptom onset, whereas the horizontal dotted lines show the threshold probability where the cumulative probability of infectiousness above the threshold is 95% of the overall cumulative probability. Colored values show the percentage presymptomatic transmission and the infectious period, i.e. the period when the viral load is above the threshold, predicted by the corresponding model. The mean durations of the infectious period across all individuals are 9.2 days and 7.5 days for *K*_*m*_ = 10^3^ and 10^4^ RNA copies, respectively. The infectious periods last on average 7.8 and 6.7 days post symptom onset for *K*_*m*_ = 10^3^ and 10^4^ RNA copies, respectively. **(I)** The relationship between the infectiousness (y-axis) and log_10_ viral load (x-axis) predicted by the probability model assuming *K*_*m*_ = 10^3^ (dark blue) or 10^4^ (green) RNA copies.

The fraction of infections that occur before symptom onset is highly dependent on *K*_*m*_ (Fig. 3 and S4). With *K*_*m*_ in the range of 10^3^ to 10^4^ RNA copies, the presymptomatic fraction of infectiousness in patients whose incubation period is between 4 and 4.5 days (as in our data) is 13%-22% (panels B, D, E, F and H in Fig. 3), a range similar to the prediction using AUClog and close to the lower bound estimated by He *et al*. (9). Further, the period of higher viral load that comprises 95% of the cumulative infectiousness, i.e. which we assume is approximately the infectious period, is on average 9.2 days (sd: ±2.5 days) and 7.5 days (sd: ±2.0 days) across the 8 patients, for *K*_*m*_ = 10^3^ and 10^4^ RNA copies, respectively (Fig. 3). This is consistent with a serial interval of 7-8 days in the absence of active tracing and isolation as estimated from Ref. (46), assuming a mean latent period, i.e. from infection to becoming infectious, of 3 days (9). The infectious periods last on average 7.9 days (sd: ±2.5 days) and 6.7 days (sd: ±2.0 days) post symptom onset, for *K*_*m*_ = 10^3^ and 10^4^ RNA copies, respectively (Fig. 3). Again, this is consistent with clinical studies showing that infectious viruses can be isolated from patient samples during the first week of symptoms and up to 8-9 days post symptom onset (5, 8, 47). Overall, the consistency between this model and epidemiological evidence suggests that *K*_*m*_ in the range of 10^3^ to 10^4^ RNA copies represents a reasonable choice (Fig. 3I).

Importantly, we found that the area under the infectiousness curve *p*(*t*) predicted by the probabilistic model, strongly correlates with AUClog calculated above (p-value<10^−10^; r^2^>0.9998), and the fractions of presymptomatic infections predicted by the probability model are very close to those predicted using AUClog (Fig. S5).

Higher values of *K*_*m*_, e.g. 10^5^ or 10^6^ RNA copies, lead to very small fractions of presymptomatic transmission that are inconsistent with data (Fig. S4). In general, when *K*_*m*_ is larger than the maximum viral load, *V*_*T*_(*t*) + *K*_*m*_ ≈ *K*_*m*_, and thus 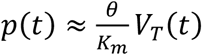.In this case, the infectiousness is directly proportional to the viral load and the fractions of presymptomatic transmissions with such a large *K*_*m*_ are close to those predicted by the %AUC (Fig. 2A), and are too small to be consistent with data. In addition, when infectiousness is proportional to AUC, most infections are predicted to occur within a few days of the viral load peak, which is inconsistent with a relatively long infectious period (5, 8, 46, 47). Therefore, including a saturation effect in viral transmission, here modeled by the Michalis-Menten term, is key to having the probability model predict a large fraction of presymptomatic transmissions.

Sensitivity analyses with respect to the value of *θ* show changes in the overall probability of transmission; however, the proportion of presymptomatic transmission and the duration of infectiousness remain largely unchanged (Figs. S6 and S7). Overall, the probabilistic model of infectiousness based on the early viral load dynamics in the URT explains our findings and is consistent with multiple epidemiological observations.

### Long-term dynamics of SARS-CoV-2 indicate continuous infection of new target cells in the LRT

Next, we analyzed data collected up to 31 days post infection. One interesting pattern we observe in the LRT is that the viral load is maintained at intermediate-to-high levels for a prolonged period of time after the first viral peak in most infected individuals. In addition, multiple viral peaks were observed in the LRT of several individuals (see Figs. 4A, B, D, E, F and G). We hypothesize that these additional peaks could be due to infection of new target cells in the LRT. These target cells can come from three potential sources. First, it is possible that after the initial viral peak, viral load decreases exponentially, and this decrease leads to decreases in the level of interferon and other cytokines and subsequent shut-down of the antiviral response. Cells in an antiviral state, which have been called cells refractory to infection or simply refractory cells (48), could then become target cells again, fueling the continued infection in the LRT. Second, cells in the LRT may proliferate and replace cells killed by the virus. Type II alveolar cells, which are the predominant target cells in the LRT, are capable of proliferating and can also differentiate into type I alveolar cells, which also can be infected by SARS-CoV-2 (19, 39). These two cell types make up over 90% of the cells in the LRT (49, 50). Third, it is plausible that infection in the LRT is highly spatial as suggested by CT scans (51, 52). Viruses may move to new physiological locations in the lungs over time and find new target cells to infect, which leads to maintenance or increases in viral load in the LRT (5).

**Figure 4.**
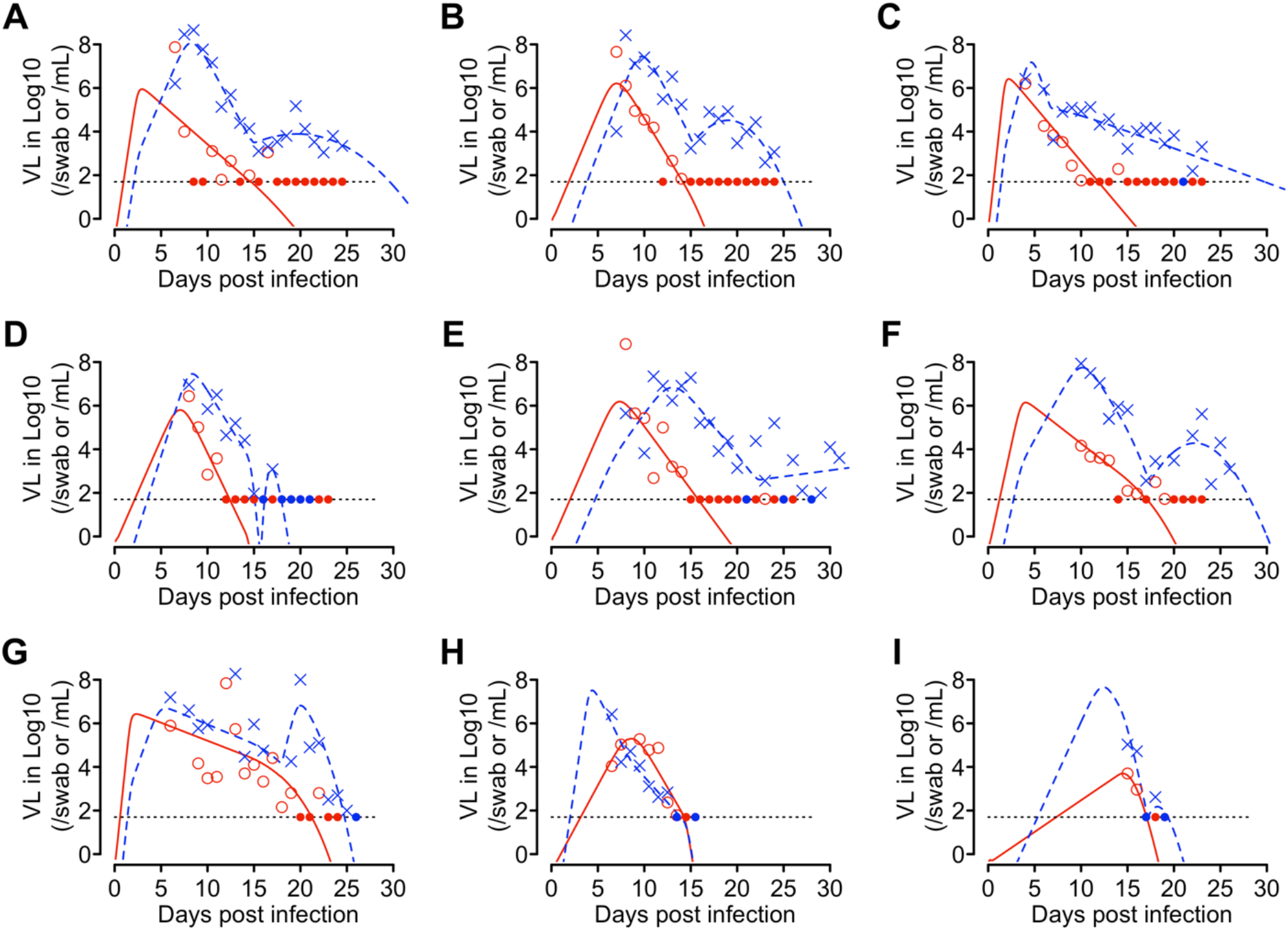
The extended target cell model describes the long-term viral load data from the URT and the LRT well. Red and blue denote viral load kinetics in the URT and the LRT, respectively. Lines denote simulations using best-fit parameter values for each individual (Table S6). Symbols show the data from throat swabs (red squares) and the sputum samples (blue ‘x’s) as reported in Ref. (5). The dashed black lines show the limit of detection, and closed dots show data below the limit of detection.

To test these hypotheses, we developed alternative models by extending the TCL model. Since it is likely that adaptive immune responses develop after 14 days of infection (38), we incorporated the impact of adaptive immune responses, following the framework of Pawelek et al. (53), in all the models we develop below to analyze long-term data. Here, we focus on the immune responses that increase the rate of infected cell killing (see Supplementary Material), such as cytotoxic T lymphocyte responses and antibody responses (54) that lead to infected cell killing via mechanisms such as antibody-dependent cellular cytotoxicity, antibody-dependent phagocytosis and complement-mediated cell death.

We tested alternative models incorporating the innate immune response (i.e. the innate immunity model), proliferation of target cells (i.e. the proliferation model), spatial spread of the virus in the lungs (i.e. the extended target cell model) or both the innate immune response and the spatial spread in the lungs (i.e. the combined model) as described in Methods. We used these models to understand the full viral load dataset encompassing viral loads measured to day 31 in Ref. (5). Because of the highly heterogeneous long-term viral load dynamics, we fitted the models to data from each individual separately (see Methods and Supplementary Material).

Fitting results (Fig. 4 and S8-S11) and model comparison using AIC scores (Table S5) show that the extended target cell model is the best model. It describes the multiple viral load peaks seen in the longitudinal sputum samples well (Fig. 4). See Table S6 and S7 for best-fit parameter values and their confidence intervals. Although the proliferation model had a higher overall AIC score than the extended target cell model, it describes the data from some patients almost as well as the extended target cell model based on the AIC scores for individual patients (e.g. Patients 2, 7, 10, 14; see Table S5). The models without the appearance of new target cells during infection (i.e. the TCL model and the innate immunity model) failed to describe the multiple viral load peaks seen in the sputum samples from many individuals (Fig. S9 and S10). Overall, the spatial infection hypothesis that viruses find new target cells to infect over the course of infection in the lungs can explain the prolonged period of intermediate-to-high levels of virus in the LRT. Proliferation of type II alveolar cells may also serve a source for new target cells to sustain viral infection in the lungs.

### Impact of therapeutics

We next used the extended target cell model to evaluate the impact of potential therapy on viral dynamics and infectiousness of each individual. We included in the model the effects of therapeutics that reduce virus production and/or therapeutics that inhibit virus entry to host cells (see Supplementary Information). Similar results were obtained with administration of each of the antivirals alone or combinations of the two (Figs. S12-S14).

For the dynamics in the URT, we focused on the first 14 days of infection, during which time infectious virus can be recovered (5). In general, we found that a potential therapeutic can reduce infectiousness by greater than 50% only when the antiviral is highly efficacious (>90%) and is administered at symptom onset (Figs. 5 and S12A-C). The maximum reduction in infectiousness among the 8 patients is predicted to be around 80% (Fig. S12A). We note that when the efficacy of the antiviral is less than 60%, the AUClog can slightly increase in some patients (see Fig. 5 and Fig. S12A-C). This is because although a suboptimal antiviral reduces the peak viral load, it may reduce the growth rate of the virus such that the viral load is maintained at intermediate levels for a longer period of time (“flattening the curve”), leading to a higher AUClog and a longer infectious period (Fig. 5). A similar effect was previously predicted in the case of antiviral therapy for Zika virus (27) and in a stochastic model of the action of prophylatic therapy to prevent SARS-CoV-2 (32). We caution that further clinical data is needed to know whether this predicted increase in AUClog has any clinical or epidemiological significance.

**Figure 5.**
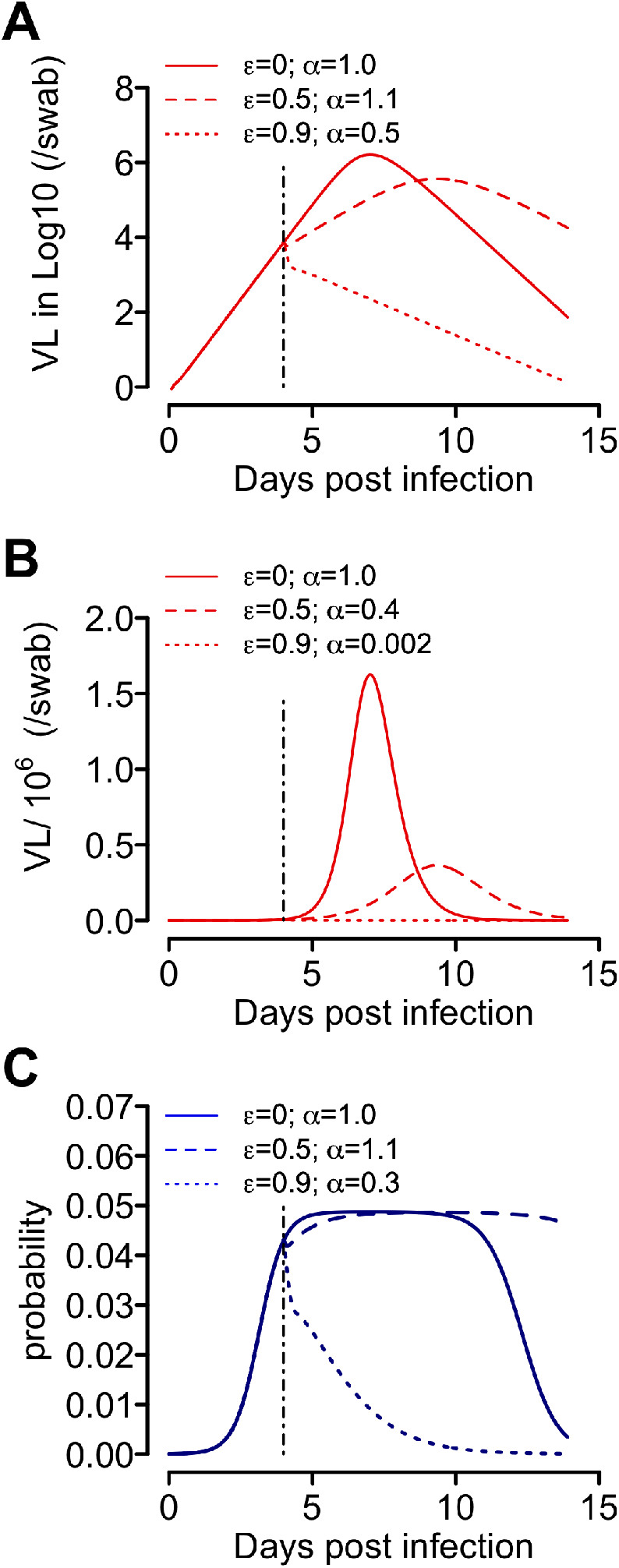
The impact of therapy on (A) the log viral load curve, (B) the viral load curve and (C) the predicted infectiousness. Solid lines show the time course simulation of the extended target cell model using best-fit parameters for Patient 2 (red) and the predicted infectiousness for the patient (blue) without therapy. Dashed lines and dotted lines show the predicted impact of a therapeutic with 50% efficacy (*ϵ* = 0.5) and a therapeutic with 90% efficacy (*ϵ* = 0.9), respectively. Both are assumed to be administered at the day of symptom onset, i.e. 4 days post infection (vertical black line). The value of *α* is the AUC of the curve of interest normalized by the AUC of the baseline curve without therapy in each panel. Thus, the value of *α* denotes the relative change of AUC compared to no treatment.

Our results highlight the importance of using the appropriate surrogate measure, i.e. AUClog, for predicting infectiousness. If AUC was used to approximate infectiousness (instead of AUClog), one would substantially overestimate the efficacy of therapy. For example, the maximum reduction in infectiousness among the 8 patients is predicted by the AUC to be ∼99.9%, instead of 80% predicted by AUClog (Fig. S12D). Further, when the efficacy of the antiviral is 50%, using the AUC one would predict substantial reduction in infectiousness, whereas using AUClog no reduction in infectiousness is observed (Figs. 5 and S12D).

For the dynamics in the LRT, we found that when administered early (i.e. at symptom onset; Fig. S15), a highly efficacious antiviral (e.g. with 90% efficacy) may change the course of infection by substantially reducing the viral load. A therapeutic with a moderate efficacy of 50%, can reduce and delay the LRT peak viral load in most patients. When administered at 8 days after symptom onset (Fig. S16), such as in the remdesivir clinical trial reported in Ref. (55), a hypothetical therapy with 50% efficacy reduces the second viral peak in some patients, but not others. In our model, the second viral peak is likely because of spread of the virus into new physiological locations in the lungs, and thus such therapy may be able to restrict the spread of the virus in the lungs in some patients. Further work is needed to quantify how disease severity is related to viral load and spatial spread in the lungs so as to be able to predict how therapeutics impact clinical outcomes.

## Discussion

In this study, we constructed mathematical models to describe the viral dynamics of SARS-CoV-2 in both the URT and the LRT, and fit the models to data collected from human subjects reported in Ref. (5). Because we used the known infection dates, we could estimate the initial dynamics more carefully than previous studies (32-36). In particular, we found infected individuals with a longer incubation period had lower rates of viral growth, and higher potential for presymptomatic transmission. We found that the logarithm of viral load in the URT may serve as a good surrogate of the infectiousness of an individual, and a physiologically based model incorporating viral shedding and establishment of infection is able to explain this relationship. Finally, we provided evidence that continuing viral infection of new target cells in the lungs may be the reason high LRT viral loads are maintained for a sustained period.

We first used a target cell limited model (26) to understand early viral dynamics (up to day 14 post infection) in the URT and the LRT. Through a population fitting approach, we estimated that the viral load peaks on average around 2 days (±0.2 day) and 2.1 days (±1.2 days) post symptom onset, respectively, in the URT and the LRT. The duration of the incubation period, i.e. the time from infection to symptom onset, is negatively correlated with the rate of early viral load increase, i.e. the quicker the virus load reaches a high level in the URT, the shorter the incubation period. This suggests that the initial symptom development may be driven by a high level of virus in the URT. These results were only possible because in this dataset the day of infection and the day of symptom onset were both available. The incubation periods of the patients we analyzed in this study are relatively short (i.e. 1-4.5 days). Studies involving a larger number of patients (with known infection dates) with incubation periods spanning longer durations (i.e. >5 days) will be useful to further inform the relationship between viral dynamics and symptom development. Moreover, longitudinal measurements of viral load at the time of or before symptom onset could help refine our estimates, but such data is unlikely to be available.

The quantitative relationship between viral load and infectiousness of an individual is key to linking within-host viral load to infectiousness and transmission dynamics (12). For SARS-CoV-2, infectiousness is likely related to viral loads in the URT (11), and a sizable fraction of transmission occurs before symptom onset (9). To explain this epidemiological observation, we developed a probabilistic model that considers viral shedding from a donor and the establishment of infection in a recipient during a contact, and show that a saturation effect, where infectiousness saturates when viral load becomes very high, is key to explain a large fraction of presymptomatic transmissions. We note that a saturation effect was also found to be important to describe how infectiousness depends on viral load in HIV infected (41) and influenza A infected individuals (42). Importantly, because of the saturation effect, we show that the logarithm of viral load (rather than absolute viral load) is an appropriate surrogate for infectiousness. Indeed, if infectiousness was directly proportional to viral load, i.e. no saturation effect, we would expect little presymptomatic transmission and a person would be infectious for only a day or two when viral load is near its peak. These predictions are inconsistent with clinical and epidemiological observations that infected persons on average are infectious for a week or so (5, 8, 46, 47).

The results of our probabilistic model and the finding that logarithm of viral load is a more appropriate surrogate measure for infectiousness than the viral load have important implications. First, as we have shown, using the viral load instead of its logarithm as a surrogate for infectiousness substantially overestimates the effectiveness of therapeutics on infectiousness. Second, there is an emerging need to quantify the extent of transmission of asymptomatically infected individuals and children (56). Our model provides an important tool for predicting infectiousness of these groups of individuals based on their viral load data (13, 14). These predictions will be useful for making public health policies, such as school reopening (57).

The probabilistic model we developed represents a first step to link viral load to SARS-CoV-2 infectiousness, but it has limitations. First, the model used a Michaelis-Menten term to model the saturation effect for simplicity. The data we used to support this functional form were limited and other functional forms incorporating a saturation effect may also be able to explain the data. Second, the model makes several simplifying assumptions including assuming that all contacts are of the same duration and the parameter values in the model are the same across patients, whereas in reality, there may exist considerable heterogeneity (43, 58). Therefore, further model developments incorporating different aspects of heterogeneity in the transmission process (40, 58), rigorous parameter estimation from clinical and epidemiological data, and testing of alternative models are warranted.

The viral load in the LRT is maintained at intermediate-to-high levels for a prolonged period of time (5, 16). In Ref. (5), multiple viral peaks in the LRT were observed in several patients. We tested different hypotheses to explain this observation by constructing mathematical models that include the immune responses, proliferation of target cells or spatial spread of virus in the lungs. The best model for explaining the data is the one that implicitly considers spatial spread of virus in the lungs. If virus can reach new physiological compartments, e.g. different regions of the lungs, then new target cells become available to fuel further infection. This finding is also consistent with data from chest CT scans of infected individuals showing that infection spreads progressively to larger areas in the lungs (51, 52). More broadly, it has been suggested that infection is highly spatial for other viruses such as influenza (59), and thus, the spatial nature of within-host virus spread may be a general feature of respiratory viruses. However, more research (similar to what has been done in influenza, e.g. Ref. (60)) is needed to investigate how viral spread in the lungs, proliferation of target cells and the immune response lead to different levels of symptom severity and disease outcome.

We found that adding a type I interferon response to the model does not improve the fit to the data. While it is possible that the resolution of the data does not allow us to detect the impact of the interferon response on viral kinetics; it is possible that the interferon response is suppressed by SARS-CoV-2 (61-63), at least early on, and thus may play a minor role in regulating viral kinetics. A recent single cell analysis of SARS-CoV-2 infection shows that SARS-CoV-2 induces low levels of type I and III interferons, but an elevated level of inflammatory chemokines (64). Whereas other studies with patient samples have indicated that the type I IFN response is more overt in severe cases of infection, i.e., later in the course of the disease (65). If this is the case *in vivo*, the lack of a potent interferon response to stop/limit virus spread early and the subsequent excessive production of inflammatory molecules as a result of continuous viral infection can lead to lung damage and severe clinical outcomes (66).

In summary, we developed a mathematical model to describe the dynamics of SARS-CoV-2 infection in both the URT and the LRT. Using this model, we provided several important quantitative estimates of the features of infection dynamics, including the number of target cells, the time to peak viral load, and the impact of viral dynamics on the length of the incubation period and the infectiousness of a host. We note that the data we used for model inference are from infected individuals with relatively mild symptoms (5). The estimated parameter values thus may be biased towards mildly symptomatic individuals. Further work is needed to compare the viral dynamics in individuals with different levels of symptom severity, e.g. asymptomatic individuals (13), individuals with severe symptoms and critically ill patients (67). Overall, this model serves as a crucial step towards a framework for better quantitative understanding of SARS-CoV-2 dynamics, the host immune response and the impact of therapeutics on infectiousness and disease symptoms of an infected individual.

## Methods

### Mechanistic models

#### Target cell limited (TCL) model

We first construct a within-host model based on the target cell limited (TCL) model used for other respiratory viruses, such as influenza (26) and respiratory syncytial virus (68). The model keeps track of the total numbers of target cells (*T*), cells in the eclipse phase of infection (*E*), i.e. infected cells not yet producing virus, productively infected cells (*I*) and viruses (*V*). The compartments in the URT and the LRT are subscripted with ‘1’ and ‘2’, respectively. To compare the model with data, we keep track of sampled viruses, i.e. virus levels measured in pharyngeal *throat* swabs, *V*_*T*_, or *sputum* samples, *V*_*S*_, and assume that these levels are proportional to the number of viruses in the URT and the LRT (*V*_1_ and *V*_2_), respectively. The ordinary differential equations (ODEs) describing the model are

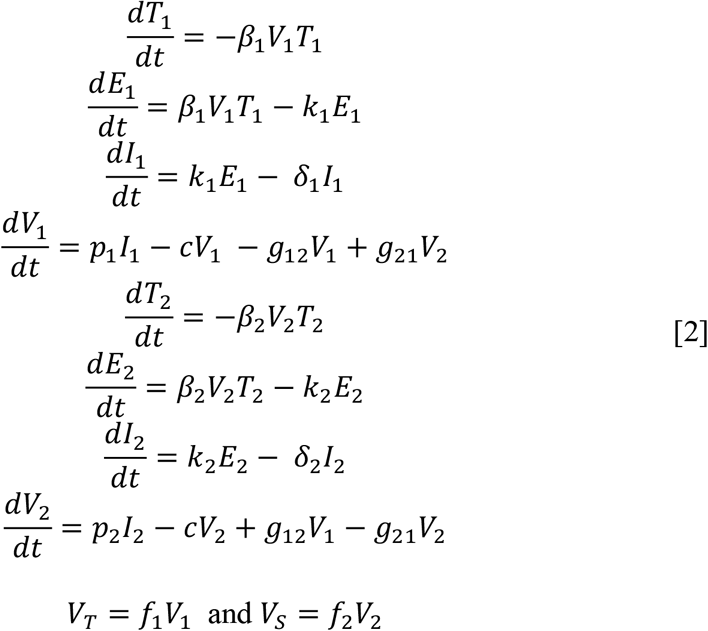

In the URT, target cells are infected by virus with rate constant *β*_1_. Cells in the eclipse phase become productively infected cells at per capita rate *k*_1_. Productively infected cells produce virus at per capita rate *p*_1_ and die at per capita rate *δ*_1_. Viruses are cleared at per capita rate *c*, and are transported from the URT to the LRT at rate *g*_12_, the opposite movement, from the LRT to the URT occurs at rate *g*_21_. The equations in the LRT follow the same model structure. *f*_1_ and *f*_2_ are the proportions of virus sampled in the two compartments, i.e., *f*_1_ = *V*_*T*_/*V*_1_ and *f*_2_ = *V*_*S*_/*V*_2_. Note that because our model keeps track of the total number of viruses and not their concentrations in the URT and LRT, the relative volumes of these compartments do not enter in the description of the transport processes between these compartments.

#### Model simplification

For practical data fitting and parameter estimation, we make several assumptions to reduce the number of parameters in the model. First, we derive ODEs for *V*_*T*_ and *V*_*S*_, i.e. the measured viral load observables, to replace the expressions for *V*_1_ and *V*_2_. Hou et al. provided evidence supporting the idea that the URT is the initial site of infection and virus is later seeded to the LRT through the oral-lung aspiration axis (69). Thus, the transport of virus from the URT to the LRT is likely to occur in random episodes, and here we approximate that by a slow continuous transport, which we assume is much slower than viral clearance, i.e. *g*_12_ ≪ *c*; thus *c* + *g*_12_ ≈ *c*. There is no evidence that the viral dynamics in the LRT have strong impact on the viral dynamics in the URT. For example, the viral load in the URT typically declines to very low levels after initial peak while the viral load in the LRT is sustained at higher levels for a long period of time (5). Thus, we assume that *g*_21_ ≈ 0. (see Supplementary Material for further justifications of these assumptions). Under these assumptions, the ODEs for *V*_*T*_ and *V*_*S*_ in this simplified TCL model become:

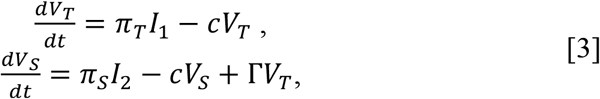

where 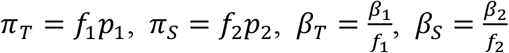 and 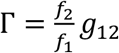. The ODEs for the host-cell compartments are unchanged.

From this model, we calculate the within-host reproductive number for the dynamics in the URT, *R*_0,*URT*_, as:

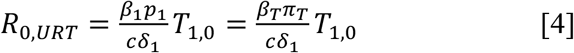

where *T*_1,0_ is the initial number of target cells in the URT.

If we assume that the number of viruses transported from the URT to the LRT is small compared to the number of viruses produced from infected cells in the LRT after initial seeding of infection in the LRT, the within-host reproductive number for the dynamics in the LRT, *R*_0,*URT*_, can be approximated as:

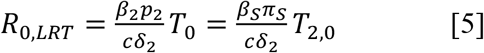

where *T*_2.0_ is the initial number of target cells in the LRT.

See Supplementary Information for the details for the choice of fixed parameter values and for other models (i.e. the innate immunity model, the proliferation model, the extended target cell model and the combined model) describing the long-term viral load dynamics.

### Probability model linking viral load in the URT to infectiousness of a host

We construct a simple model linking within-host viral load in the URT in a donor to the probability of transmission to a recipient. We consider a typical contact of length *τ* between the donor and the recipient at time *t*. We assume that *τ* is small enough (on the order of minutes or hours) that the viral load in the URT of the donor *V*_1_(*t*), and thus *V*_*T*_(*t*) is approximately constant during the contact. A recent experiment showed a positive correlation between the number of seasonal coronaviruses in a throat swab and the number of viruses in respiratory droplets or aerosols (11). However, the number of virus shed in respiratory droplets or aerosols appears to saturate when the viral load in a throat swab becomes high, e.g. exceeding 10^4^ copies (Fig. S3). Thus, we used a Michaelis-Menten term to model the amount of virus shed from the host’s URT: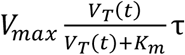, where *V*_*max*_ is the maximum viral particles shed from a host per unit time, and *K*_*m*_ is the saturation constant. When *K*_*m*_ is much greater than the peak viral load, saturation will not be observed, so this model incorporates both a linear and saturating dependence on viral load.

Next, we assume that a fraction of the shed virus, *φ*, reaches the URT of the recipient and that a virus that reaches the recipient’s URT has a probability *v* to successfully establish infection. Then, the number of viruses that successfully establish an infection is given by a binomial distribution Bin(*n*; *v*), where 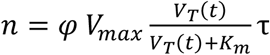. For large *n* and small *v*, Bin(*n*; *v*) is well-approximated by a Poisson distribution with parameter 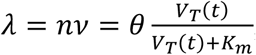, where *θ* = *vφ*τ *V*_*max*_. The probability of one or more virions generating a successful transmission event for a typical contact at time *t* is *p*(*t*)=1-exp(-λ) as shown in Results.

### Data, parameter fitting, analysis and model selection

We digitalized longitudinal viral load data from throat swabs and sputum samples of the 9 infected individuals reported in Wolfel et al. (5). The infected individuals are young to middle-aged professionals, without underlying disease, who were identified because of known close contact with an index case. All patients were hospitalized, but had a comparatively mild clinical course of disease.

We took two approaches for model fitting. First, when fitting models to data collected up to day 14 post-infection, we use a population approach, based on non-linear mixed effect modeling, to fit the model simultaneously to viral load data from the URT and LRT of 8 patients. Second, when fitting models to long-term time series involving all data collected up to day 31 post infection, we fit the model to data from each patient separately. This approach is taken due to the substantial variability in the long-term viral dynamics among individuals, which precludes us from assuming they are a sample from a homogeneous population. See supplementary text for details of these two parameter fitting procedures, including parameters estimated and parameters kept fixed with sensitivity analyses. We calculated correlations between viral characteristics parameters and the incubation periods using Pearson correlation.

To compare the different models tested, we calculate Akaike Information Criteria (AIC) scores from the residual sum of squares (RSS) as

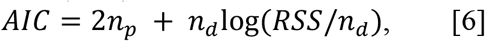

where *n*_*p*_ is the number of fitted parameters and *n*_*d*_ is the number of data points used in estimation (70).

## Data Availability

The data are available in the manuscript and the supplementary materials.

## Acknowledgements

Portions of this work were done under the auspices of the U.S. Department of Energy through Los Alamos National Laboratory, which is operated by Triad National Security, LLC, for the National Nuclear Security Administration of the U.S. Department of Energy (contract No. 89233218CNA000001). The work was supported by the Laboratory Directed Research and Development program of Los Alamos National Laboratory (project No. 20200743ER and 20200695ER), and by the Defense Advanced Research Projects Agency (contract No. HR0011938513). Part of this research was supported by the DOE Office of Science through the National Virtual Biotechnology Laboratory, a consortium of DOE national laboratories focused on response to COVID-19, with funding provided by the Coronavirus CARES Act.

